# Biostatistical Investigation of Correlation Between COVID-19 and Diabetes Mellitus

**DOI:** 10.1101/2020.11.21.20235853

**Authors:** Zahra Eynizadeh, Zahra Ameli, Mahdieh Sahranavard, Mobina Daneshparvar, Mobina Abdollahi Dolagh, Mahboobeh Roozkhosh, Mohammad Reza Ejtehadi, Maryam Azimzadeh Irani

## Abstract

COVID-19 is a highly infectious disease. Studies suggest that its severity is amplified in patients diagnosed with Diabetes Mellitus. In this study, the correlation between the prevalence of COVID-19 and Diabetes was analyzed at the regional and global scale using data extracted from WHO and IDF Diabetes Atlas. For the regional investigation data was assorted into ten regions including Central Asia, Middle east and western Asia, Africa, North America and the Caribbean, South east Asia, East Asia, Europe, South and Central America, South Asia and Oceania. The results show a positive correlation coefficient of 0.47 in Middle east and western Asia. While at the global scale analysis all the selected countries were considered together and a correlation coefficient of 0.32 was observed. This number was increased to 0.69 when the top most affected countries by COVID-19 were considered for the analysis. In order to investigate the time dependent relationship of the two diseases, the data was analysed in five windows of 45 days each since the beginning of pandemic. The results show an increasing pattern of the correlation coefficient in the last three windows. Overall, based on this study by increasing the prevalence of Diabetes Mellitus, the prevalence of COVID-19 cases may also increase.

## 1. Introduction

Following the occurrence of atypical pneumonia in Wuhan China, and subsequently in Thailand, Japan, South Korea, and eventually the rest of the world, a pandemic was announced by the World Health Organization (WHO) on 11th of March 2020. The first infected human cases were reported on December 19th 2019 [1]. The disease was extensively spread around the world and to this date more than 40 million cases of COVID-19 have been reported and over 1.1 million patients have died.

This is the third pandemic caused by beta coronaviruses in the last two decades; the Severe Acute Respiratory Syndrome Coronavirus (SARS-CoV-1) emerged in 2002, with a mortality rate of 10% and the Middle East Respiratory Syndrome Coronavirus (MERS-CoV) emerged in 2012 with a mortality rate of 35% [2, 3].The coronavirus disease 2019 (COVID-19) is caused by the Acute Respiratory Syndrome Coronavirus 2 (SARS-CoV-2), the origin of which is believed to be bats as the primary hosts [4] and the Malayan pangolin as the intermediate host [5]. The virus is extremely successful in hijacking the human cells and presents a high infection rate.

Social distancing, home quarantines, and travel restrictions are the main strategies in the management of the disease. Due to the uneven spread of COVID-19 in countries, some countries have had higher numbers of COVID-19 patients while others showed a much lower prevalence.

Enormous funding has been put into making vaccines, and researchers worldwide have been looking into possible treatment options, but to this date, no cure has been found yet [6]. Recent studies indicate that among those suffering from COVID-19, patients with diagnosed Diabetes Mellitus (DM) show higher disease severity and risk of mortality [7-10]. This could be due to hypertension, cardiovascular diseases, obesity, and chronic kidney diseases often observed in diabetic patients [11].

This study puts together all the available statistical data to explore the relationship of DM and COVID-19 on a local and global scale. These analyses have immediate benefits in management and treatment of the DM patients who are affected by COVID-19.

## 2. Materials and Methods

### 2.1 Studied populations

Based on the World Health Organization and IDF Atlas of Diabetes data sets, regional and accumulative populations were chosen [12, 13]. It should be noted that some countries were omitted due to the lack of data. The regional populations were classified into 10 groups including; Central Asia, Middles east and western Asia, Africa, North America and the Caribbean, South east Asia, East Asia, Europe, South and Central America, South Asia and Oceania. Detailed list of the countries in each region is mentioned in Table 1. The accumulative population consists of all 190 countries which are mentioned in Table 1.

**Table 1.** Detailed list of countries in each of the 10 studied regions.

| Region | Countries |
| --- | --- |
| **Central Asia** | Kazakhstan , Kyrgyz Republic , Uzbekistan, Tajikistan |
| **Middle East and Western Asia** | Armenia, Azerbaijan, Bahrain, Cyprus, Georgia, Iran, Iraq, Israel, Jordan, Kuwait, Lebanon, Oman Palestine, Qatar , Saudi Arabia, Turkey, United Arab Emirates, Yemen |
| **South East Asia** | Cambodia, Indonesia, Malaysia, Myanmar, Philippines, Singapore, Thailand, ThailandTimor-Leste, Brunei Darussalam |
| **North America and Caribbean** | Mexico, Canada, United States, Jamaica, British Virgin Islands, Trinidad and Tobago, Haiti, Bermuda, Suriname, Guyana, Belize, St Kitts & Nevis, Grenada, Barbados, Sint Maarten, Dominica, Aruba, Saint Lucia, Curaçao, Antigua and Barbuda, Montserrat, Cayman islands, Anguilla, Greenland, Bahamas, Saint Vincent and the Grenadines |
| **Africa** | Nigeria, Ethiopia, Egypt, DR Congo, Tanzania, South Africa, Kenya, Uganda, Algeria, Sudan, Morocco, Angola, Ghana, Madagascar, Cameroon, Côte d'Ivoire, Burkina Faso, Mali, Malawi, Zambia, Senegal, Zimbabwe, Guinea, Rwanda, Burundi, Tunisia, Togo, Libya, Congo, Eritrea, Gambia, Botswana, Lesotho, Eswatini, Seychelles, Benin, Cabo Verde, Central African Republic, Chad, Comoros, Djibouti, Equatorial Guinea, Gabon, Guinea-Bissau, Liberia, Mauritania, Mauritius, Mozambique, Namibia, Niger, Sao Tome and Principe, Sierra Leone, Somalia, South Sudan |
| **Oceania** | Australia, Papua New Guinea, New Zealand, Fiji, New Caledonia, French Polynesia, Guam |
| **East Asia** | China, Japan, South Korea, Mongolia |
| **Europe** | Russia, Germany, France, Italy, Spain, Ukraine, Poland, Romania, Belgium, Czechia, Greece, Portugal, Sweden, Hungary, Belarus, Austria, Serbia, Switzerland, Bulgaria, Finland, Slovakia, Norway, Croatia, Republic of Moldova, Bosnia and Herzegovina, Albania Lithuania, North Macedonia, Slovenia, Latvia, Estonia, Luxembourg, The United Kingdom, Netherlands, Ireland, Andorra, Monaco, Liechtenstein, San Marino, Malta , Iceland , Faroe Islands |
| **South and Central America** | Argentina, Cuba, Nicaragua, Bolivia, Dominican, Paraguay, Brazil, Ecuador, Peru, Chile, El Salvador Puerto Rico, Colombia, Guatemala, Uruguay, Costa Rica, Honduras, Venezuela, Panama |
| **South Asia** | India, Pakistan, Afghanistan, Bangladesh, Nepal, Sri Lanka, Maldives |

### 2.2 Research Variables

In order to study the relationship between the number of COVID-19 and DM patients, Variables ***C*** and ***D*** were calculated; where ***C*** is the prevalence of COVID-19 among the total population of each country (Equation. 1) and ***D*** is the prevalence of DM in adults (ages 20 – 79) of the total population in each country (Equation. 2). Calculation of ***C*** and ***D*** variables was carried out by Microsoft Excel 2016.

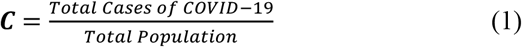

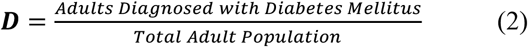

### 2.3 Data Collection

The number of adults who were diagnosed with DM for each country was taken from IDF Diabetes Atlas (https://www.diabetesatlas.org/en/data/). The data was last updated 25th of November 2019.

The total population of the countries was extracted from Worldometers (https://www.worldometers.info/world-population/). The number of confirmed cases of COVID-19 was taken from WHO webpage (https://covid19.who.int/). This data was last updated on 11th of October 2020.

### 2.4 Test of normality

The collected data for both ***C*** and ***D*** variables were evaluated to check whether they present a normal distribution. The normality of the data was tested using the Shapiro-Wilk test of normality via IBM SPSS Statistics 26 [14] which is widely used [15-18]

### 2.5 Calculation of the Correlation Coefficient

To assess the statistical relationship between ***C*** and ***D***, the correlation coefficient between the two variables, was calculated using Minitab 19 Statistical Software [19]. Among the studied populations, if both variables were normally distributed, Pearson correlation coefficient and if normality was not met for one or both variables, Spearman’s rank correlation coefficient was calculated [20].

Notice that the correlation does not necessarily indicate that a cause-and-effect relationship is at work.

### 2.6. Statistical tests and analyses

Microsoft Excel 2016, Minitab 19 Statistical Software [19] and IBM SPSS Statistics [14] were used to analyse the data.

### 2.7 Graphs

The scatter plots were drawn using Minitab 19 Statistical Software [19]. Graphs and tables were redesigned with Microsoft PowerPoint 2016.

### 2.8 Selective analysis of the countries with the highest rate of COVID-19

For an accurate analysis of the correlation between COVID-19 and DM, 15 countries with the highest prevalence of COVID-19 were chosen. These countries are as follows: Qatar, Bahrain, Aruba, Andorra, Israel, Panama, Kuwait, Peru, Chile, Brazil, The United States, San Marino, Oman, Maldives and Argentina.

Note that when testing the top 14 or 16 countries with the highest prevalence of COVID-19, similar results were observed.

### 2.9 Milestoning of the accumulative data

To determine the trend of ***C*** and ***D*** correlation throughout the pandemic, the data of variable ***C*** among the accumulative population were collected in five milestones with a duration of 45 days each. The start date was on 1/3/2020 and it ended on 11/10/2020. Exact dates of each window can be found in Table 2. Afterwards, the correlation coefficient between ***C*** and ***D*** for each milestone was calculated using the method mentioned in the “Normalization and Calculation of the Correlation Coefficient between ***C*** and ***D***” section.

**Table 2.** The calculation method for milestones.

| Variable | Calculation method |
| --- | --- |
| **Mile Stone 1** | (COVID-19 Confirmed cases from 1<sup>th</sup> March - 14<sup>th</sup> April ) / Total population |
| **Mile Stone 2** | (COVID-19 Confirmed cases from 14<sup>th</sup> April - 29<sup>th</sup> May )/Total population |
| **Mile Stone 3** | (COVID-19 Confirmed cases from 29<sup>th</sup> May - 13<sup>th</sup> July )/Total population |
| **Mile Stone 4** | (COVID-19 Confirmed cases from 13<sup>th</sup> July - 27<sup>th</sup> August )/Total population |
| **Mile Stone 5** | (COVID-19 Confirmed cases from 27<sup>th</sup> August - 11<sup>th</sup> October )/Total population |

## 3. Results

### 3.1 Regional correlation between variables C and D

Correlation between two variables ***C*** and ***D*** was investigated in all regions. The results for each region is discussed in Table 3.

**Table 3.**
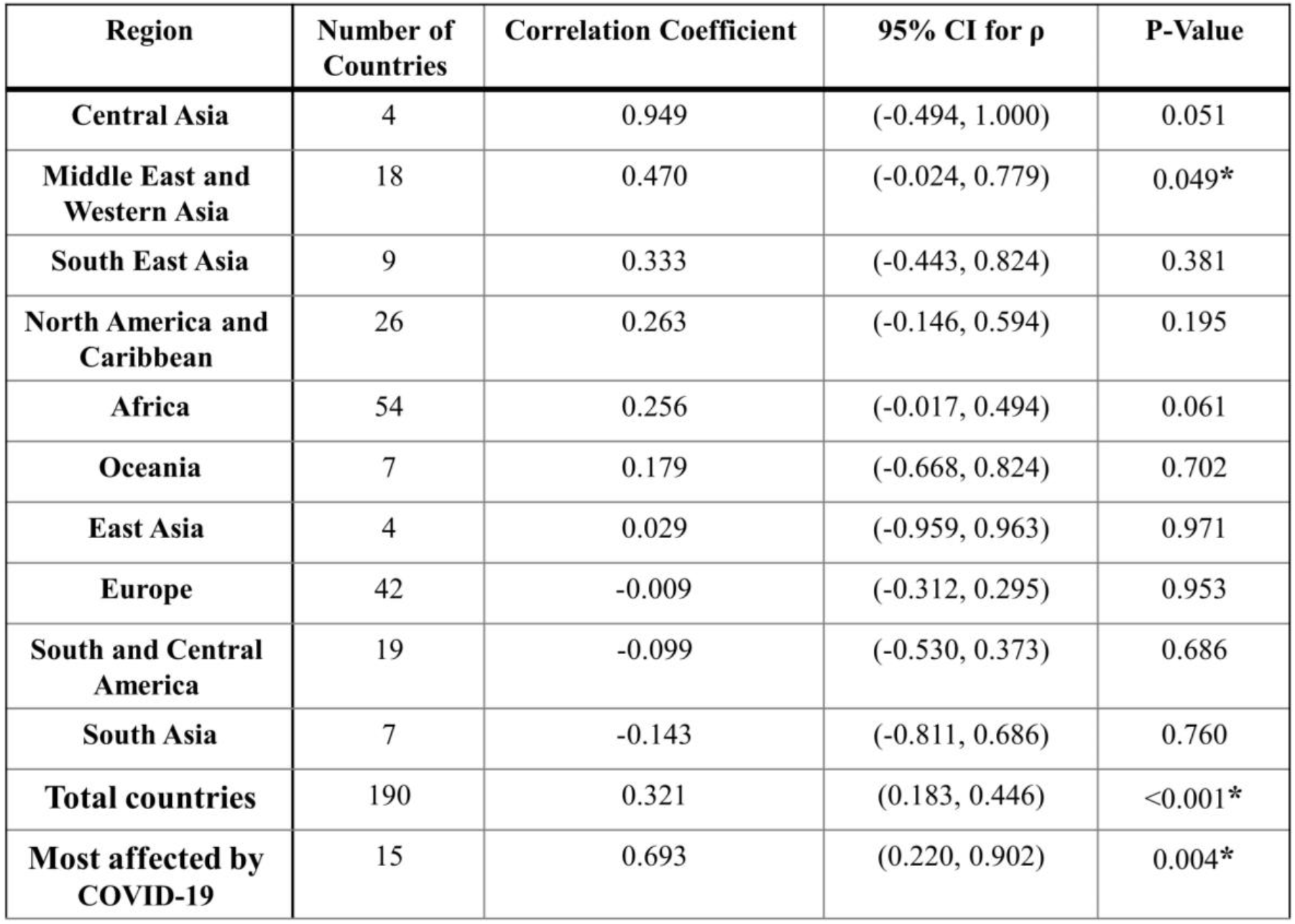
The results of statistical analysis

### 3.2 Middle East and Western Asia

In this region, the correlation between the two variables ***C*** and ***D*** was studied in 18 countries (Table 1). As shown in Table 3, the p-value is 0.049, which indicates that the correlation coefficient of 0.47 in this region is statistically significant. The positive correlation coefficient indicates that when ***D*** increases, ***C*** also increases.

### 3.3 Central Asia, South East Asia, North America and the Caribbean, Africa, Oceania, East Asia, Europe, South and Central America and South Asia

in these regions, the correlation between the two variables ***C*** and ***D*** was studied by the assortment of the countries according to Table 1. As shown in Table 3, the p-values of the calculated correlation coefficients for all of these regions are not statistically significant.

### 3.4 Accumulative Analysis

The correlation coefficient between ***C*** and ***D*** in all countries investigated in this study is statistically significant; P-Value < 0.0001. This moderately positive correlation indicates that when ***D*** increases, ***C*** also tends to increase. (Table 3 and Figure 1)

**Figure. 1.**
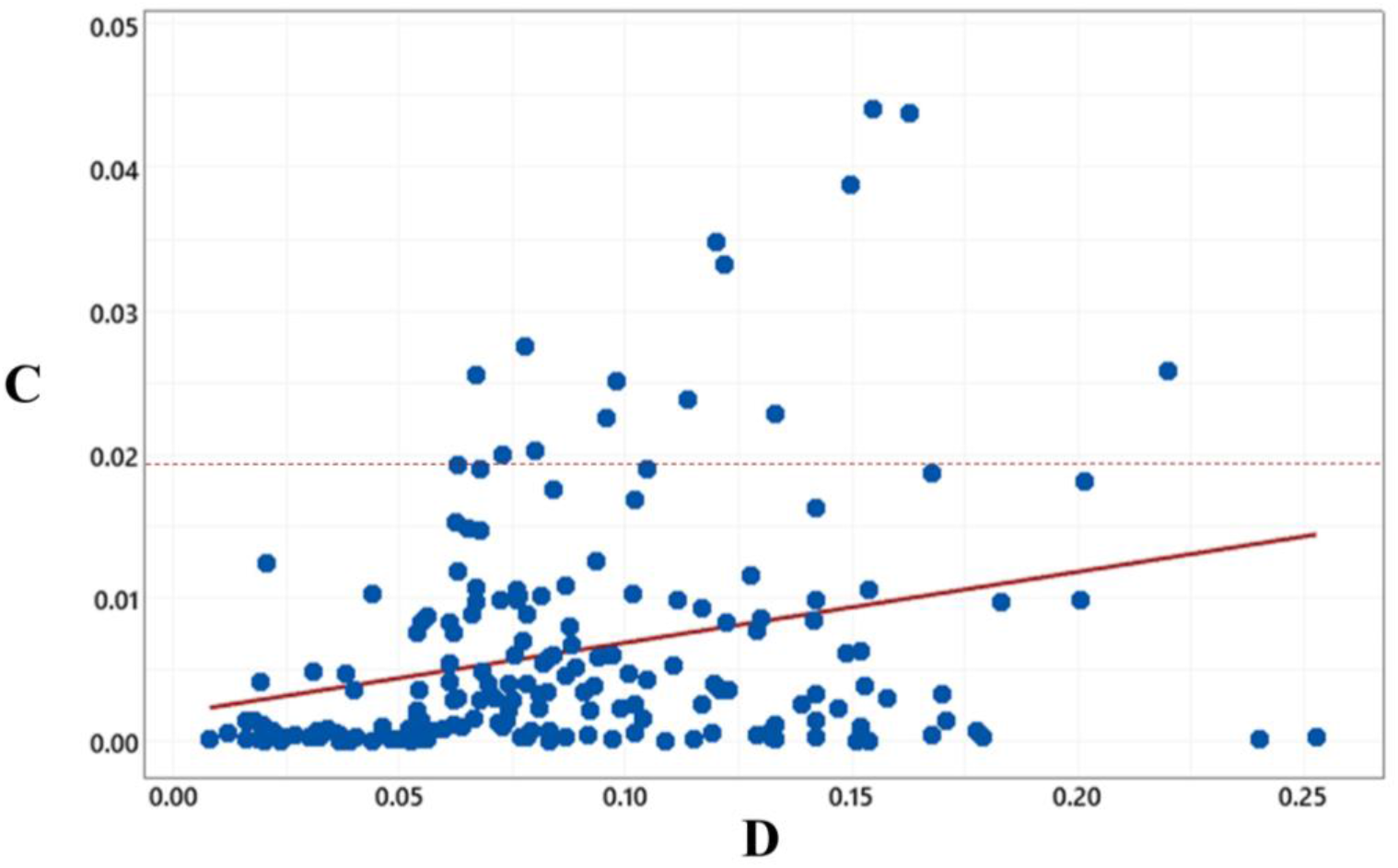
Scatterplot for C and D. An uphill pattern can be observed, this indicates a positive relationship between D and C. The dashed line shows the cut-off point for choosing the countries with the highest rate of COVID-19.

### 3.5 C and D correlation in the countries with the highest rate of COVID-19

The correlation coefficient between ***C*** and ***D*** in the 15 countries with the highest COVID-19 rate (see methods section) equals to 0.693 with P-Value= 0.018. Which indicates that there is a strongly positive correlation between the two variables (Table 3), (Figure 1 and Figure 2).

**Figure. 2.**
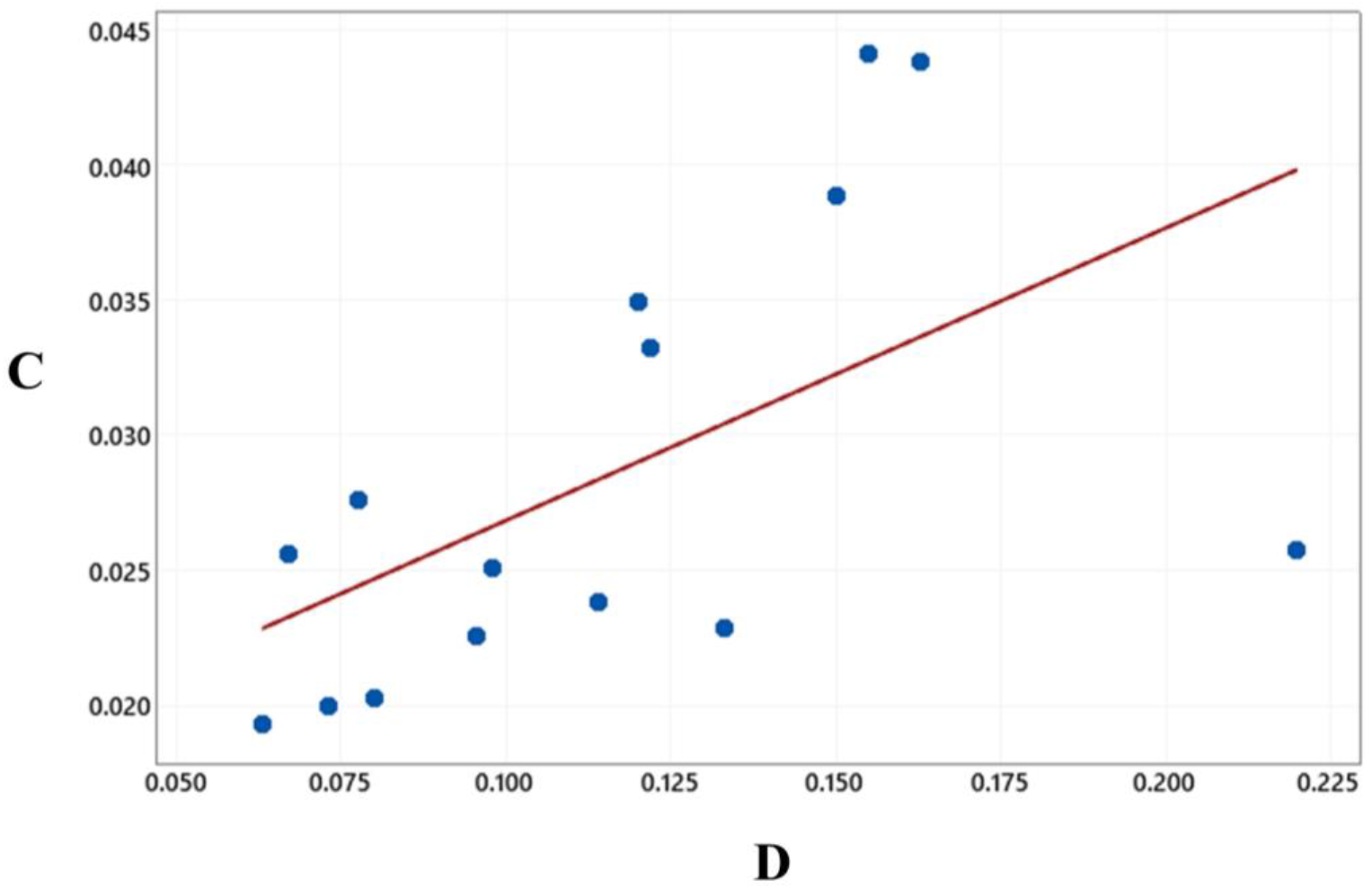
Scatterplot for C and D in top 15 countries with prevalence of COVID-19. An uphill pattern can be observed, this indicates a positive relationship between D and C.

It is worth mentioning that the reported data on the confirmed cases of COVID-19 may not be accurate in several countries. Hence, this analysis suggests that if there will be sufficient statistics on the COVID-19 cases, there could be a strong positive correlation between the two diseases.

### 3.6 Mile stones

The accumulative data was mile-stoned in windows of 45 days since the pandemic has started (Table 2). The correlation between each mile stone and ***D*** variable, indicates a significant and positive relationship in the first, second, fourth and fifth windows (Table 4).

**Table 4.** The results of statistical analysis of milestoning.

| | Number of countries | Correlation Coefficient ($\rho$) | 95% CI for $\rho$ | P-Value |
| --- | --- | --- | --- | --- |
| **Window 1** | 187 | 0.495 | (0.371, 0.602) | <0.001* |
| **Window 2** | 190 | 0.186 | (0.044, 0.321) | 0.010* |
| **Window 3** | 190 | 0.094 | (-0.049, 0.234) | 0.196 |
| **Window 4** | 190 | 0.201 | (0.059, 0.335) | 0.006* |
| **Window 5** | 190 | 0.306 | (0.168, 0.432) | <0.001* |

## 4. Discussion

This study explored the correlation between Diabetes Mellitus and COVID-19 using all the available online data until 11th of October 2020 at the local and global scale. The results show a meaningful positive correlation coefficient between the two diseases globally and in the Middle East and Western Asia on the local scale. In the countries with the highest rate of COVID-19, the correlation coefficient between the two diseases was found to be strongly positive and significantly higher than those in the accumulative study at the global scale. Overall, the results of this work indicate that if the prevalence of Diabetes Mellitus increases, the prevalence of COVID-19 cases may also increase. According to the milestoning investigation, there is an increasing pattern of the correlation coefficient in the last three windows. This increasing pattern was not observed in first and second windows, due to the uneven spread of COVID-19 and asynchronous pandemic peaks in different countries. This study brings light to the potential treatment in patients with a background of hyperglycemia and diabetes and provides a foundation for further research on the nutritional and genetics causes of the relationship between the two diseases. These findings must be expanded by paying utmost attention to collection of data from patients with hyperglycemia, DM types I and II separately and within different age groups.

With many researchers working tirelessly, every day we are getting closer to finding a vaccine for this disease. By distributing this vaccine world-wide, millions of lives will be saved. However, understanding the relationship between Diabetes Mellitus and COVID-19 as a viral infection will still remain a significant topic of research that should be expanded.

## Data Availability

IDF

## Acknowledgement

We would like to appreciate all those who have supported the production of the IDF Diabetes Atlas, 9^th^ edition, by providing additional information.

